# Label-Free SARS-CoV-2 Detection on Flexible Substrates

**DOI:** 10.1101/2021.10.29.21265683

**Authors:** Debadrita Paria, Kam Sang Kwok, Piyush Raj, Peng Zheng, David H. Gracias, Ishan Barman

## Abstract

One of the most important strategies for mitigation and managing pandemics is widespread, rapid and inexpensive testing and isolation of infected patients. In this study, we demonstrate large area, label-free, and rapid testing sensor platforms fabricated on both rigid and flexible substrates for fast and accurate detection of SARS-CoV-2. SERS enhancing metal insulator metal (MIM) nanostructures are modeled using finite element simulations and then fabricated using nanoimprint lithography (NIL) and transfer printing. The SERS signal of various viral samples, including spiked saliva, was analyzed using machine learning classifiers. We observe that our approach can obtain the test results typically within 25 minutes with a detection accuracy of at least 83% for the viral samples. We envision that this approach which features large area nanopatterning, fabrication in both rigid and flexible formats for wearables, SERS spectroscopy and machine learning can enable new types of rapid, label-free biosensors for screening pathogens and managing current and future pandemics.

## Introduction

Severe acute respiratory syndrome coronavirus 2 (SARS-CoV-2), a betacoronavirus belonging to the family of *Coronaviridae*, is implicated in the current COVID-19 pandemic. While a significant effort has been directed towards vaccination, prevention and treatment of the disease, the importance of rapid diagnosis is still important for public health strategies for future variants. It is now well known that an infected person is contagious 2-7 days prior to the onset of any symptom (*1*), emphasizing the importance of surveillance and early detection. Moreover, a rapidly mutating (*2*) virus-like the SARS-CoV-2 requires a versatile detection scheme capable of detecting a broad range of mutant strains. Thus, a rapid, inexpensive, widely deployable, and label-free diagnostic platform with a high degree of sensitivity is required for managing current and future pandemics.

The currently available detection scheme can be broadly classified into three categories - genomic sequencing, serological tests, and antigen tests. The two popular methods for detection of the viral RNA include the well-known reverse transcription real-time quantitative polymerase chain reaction (RT-qPCR) (*3*) and a more recently explored CRISPR-based test (CRISPR = Clustered Regularly Interspaced Short Palindromic Repeats) (*4*). Although highly accurate, these techniques are slow, labor-intensive, and their reliability depends on factors like sample handling, storage, transportation, and operator expertise (*5*). Moreover, the scale-up of these techniques is constrained by global supply challenges, given the huge demand for the PCR primers. ELISA (Enzyme-linked immunosorbent assay) and lateral flow immunoassays, which detects the antibody corresponding to specific viral antigens (spike protein or nucleocapsid protein in case of SARS-CoV-2), on the other hand, has low sensitivity (*6*) depending on the stage of infection, requires extensive sample preparation (*6*) and a has false positive outcome of 5-11% (*7*). Antigen tests on the other hand suffer from issues related to sensitivity and false-negative results requiring further confirmation (*8*). Thus, current virus testing methods do not offer solutions for population-wide testing due to limitations in sensitivity, intricate sample processing methods, and challenging specimen storage/transportation requirements.

Considerable efforts have been directed towards the development of various biosensors for the detection of viruses (*9, 10*). But most of the sensors require functionalizing the detector probe with antibodies specific to a virus, making them unsuitable for identifying other kinds of viruses or mutants. Moreover, knowledge about predefined labels required for the design of such sensors is a huge disadvantage for surveillance of new emerging viruses. Raman spectroscopy, which relies on the inelastic scattering of light to quantify the unique vibrational modes of molecules, has enabled accurate label-free fingerprinting of individual viral components (*11*). Moreover, structural and chemical alterations in the genome and the capsid proteins are reflected as changes in the vibrational features (*11, 12*), as shown by a prior investigation that reported the distinguishable Raman signatures of intact, intermediate and disrupted echovirus 1 particle (*12*).

However, spontaneous Raman scattering does not provide the necessary sensitivity to detect a low viral titer. Surface-enhanced Raman spectroscopy (SERS) (*13*), which intensifies a weak Raman signal arising from biological samples adsorbed on noble metal nanostructures, combines the high molecular specificity with near single-molecule sensitivity (*14*) and enables spectroscopic quantification of multiple pathogen concentrations in small volumes (*15*). An important point to consider is that, even though adding metal nanostructures enables amplification of the Raman scattering signals, similar composition of viral transport media like saliva, nasal swab or cells used to culture viruses can lead to interference with the actual viral signature. Thus, further analysis using machine learning enables a more robust, sensitive and specific detection scheme removing hindrance from the presence of unwanted biological signatures (*16–18*).

Here, we present a new platform for ultrasensitive and rapid detection of SARS-CoV-2 by exploiting SERS signatures recorded on highly reproducible plasmonically active nanopatterned rigid and flexible substrates in a label-free manner. For enhancement of the weak Raman signal from the SARS-CoV-2 virus, we develop novel nanomanufacturing paradigms for large-area rigid and flexible SERS substrates patterned by nanoimprint lithography (NIL) coupled with transfer printing. The plasmonic nanostructures are composed of a Field-Enhancing Metal-Insulator Antenna (FEMIA) architecture, with multiple alternate stacks of silver and silica (similar to metal-insulator-metal plasmonic nanoantennas). Such an arrangement has its primary resonance frequency close to the laser excitation ensuring maximum Raman signal amplification. The entire scheme of our detection technique is summarized in Figure **1**. With an experimental limit of detection of 500 nM, we were able to directly read strong signatures of the viral fusion protein of SARS-CoV-2 and H1N1 in the purified form from the SERS spectra. Further, using principal component analysis (PCA) and random forest classification we could identify four different kinds of “enveloped” RNA viruses with an accuracy greater than 83%. Using our platform on a flexible substrate, we were able to detect SARS-CoV-2 in a complex body fluid like saliva. Sensing on a flexible FEMIA substrate allows mounting the sensor on curved and flexible surfaces and wearables for rapid identification of the virus in a variety of situations.

**Figure 1:**
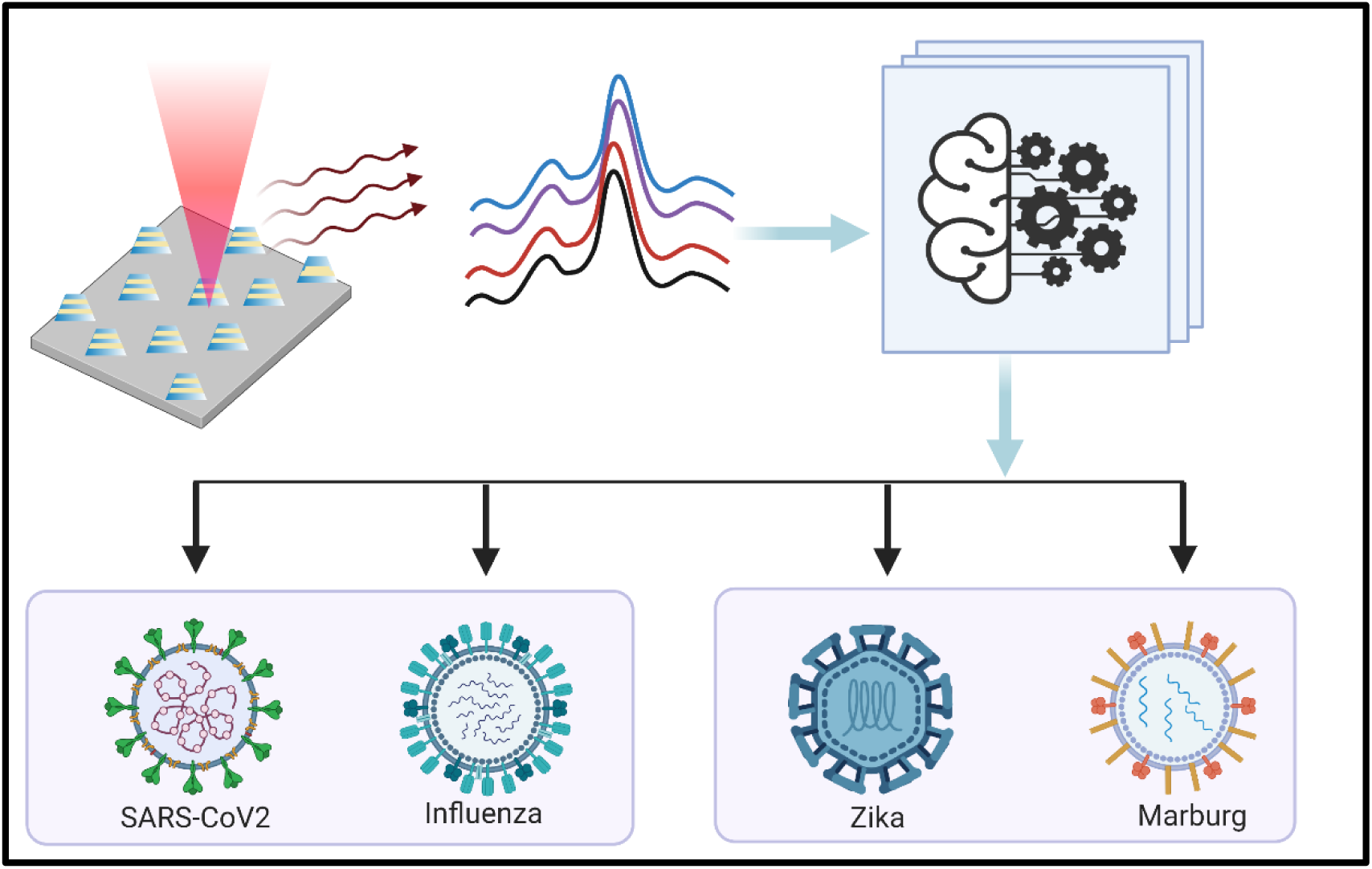
Summary of the detection scheme. SERS signals are collected from samples consisting of different types of respiratory and non-respiratory viruses placed on a nano manufactured 2D array of FEMIA. PCA and random forest classification applied on the SERS spectra allow us to distinguish and identify different viral samples.

## Results

### Fabrication of FEMIA on rigid and flexible substrate

Chemically synthesized SERS probes are plagued by irreproducible enhancement and variable spatial distributions of ‘hotspots’, leading to “SERS-uncertainty principle” (*19*). Thus, precisely fabricated plasmonic nanostructures, arranged in a regular fashion with well-defined periodicity and predictable spatial localization of near-field enhancement is the key for achieving a reproducible and measurable SERS signal. Such well-defined nanostructures can be achieved using advanced nanofabrication techniques like X-ray lithography, ion-beam lithography, extreme UV/deep UV lithography, etc., but are often expensive and require sophisticated instrumentation. Other existing methods to prepare plasmonic nanostructures for SERS substrate include bottom-up synthesis, template-assisted growth and lithography (e.g. soft lithography, nanosphere lithography and e-beam lithography), but these methods are limited by reproducibility, sensitivity, scalability and cost (*20, 21*). In contrast, nanoimprint lithography (NIL) creates a large area of well-defined nanopatterns in a parallel fashion, with a high degree of reproducibility. Also, NIL can be applied with rigid stamps or molds and can be further developed into a high throughput manufacturing process, such as roll-to-plate (R2P) or roll-to-roll (R2R) NIL process (*22, 23*).

First, we simulated the FEMIA to assess the near-field enhancing capability of the nanostructure and select the most optimum number of metal and silica layers for testing of the virus samples. A 3D geometry enables the location of the ‘hotspots’ to be raised above the substrate allowing the localized near-field to be accessible, leading to a larger interaction with the analyte molecule (*24, 25*). Along with matching the excitation to the plasmon resonance frequency of the nanostructure, a careful design to allow maximum interaction between nanostructures is necessary to achieve a significant level of SERS sensitivity. We carried out electromagnetic simulation using finite element analysis (details in Materials and Methods) to see the location of the hotspots and determine an estimate of the resonance properties of the nanostructure. Figure **2A(i)** plots the near field distribution (log_10_|E/Eo|^2^; E is the total electric field, Eo is the excitation) near the dipolar resonance (780 nm) of the FEMIA nanostructure (scheme of the simulation is provided in the Supporting Information, Figure **S1**). Enhanced near-field is observed at the corners and the sidewall of the FEMIA near the junction of the silver and the silica layer. Except for the bottom corners, where the nanostructure lies on a substrate, the rest of the spatial locations of the electromagnetic near-field are accessible to the analyte molecule. A closer look at the plasmon resonance positions for the nanostructures in the visible range reveals two dominant peaks for the FEMIA composed of 5 layers and 7 layers, whereas a single layer and 3-layer FEMIA has a single dominant resonance (Supporting Information Figure **S2**). A plot of the charge distribution of the 5-layer FEMIA reveals a higher energy multipolar resonance (Figure **2A(ii**) left panel) and a lower energy dipolar resonance (Figure **2A(ii**) right panel). The near IR dipolar resonance for the FEMIA is close to the laser wavelength used for SERS in our study.

**Figure 2:**
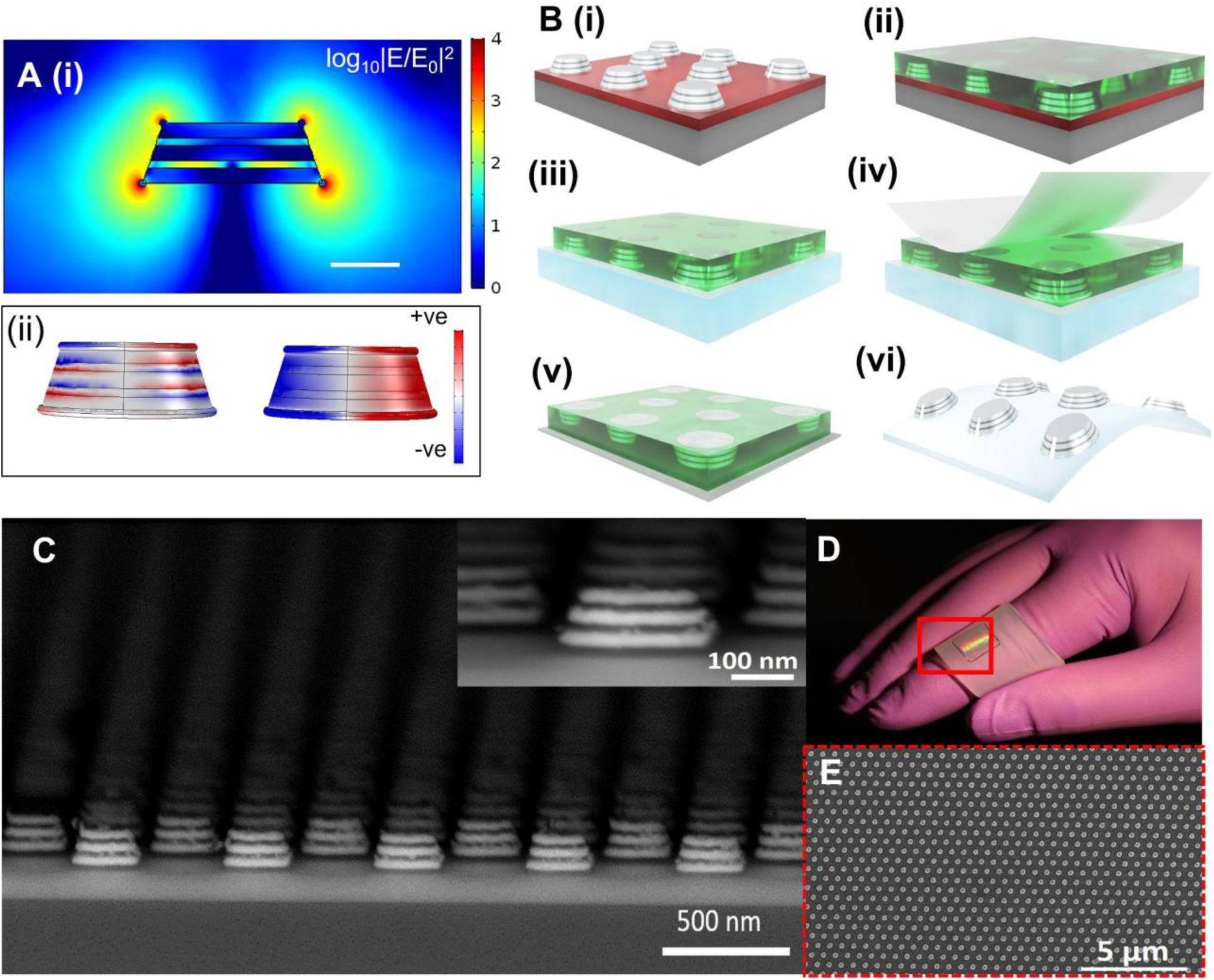
Overview of the sensor substrate fabrication. (A) (i) Plot of electric field enhancement of a simulated 5-layer FEMIA nanostructure (at 780 nm; near to the laser frequency used in the experiment) shows increased near field at the corners and the side wall. Scale bar 100 nm. (ii) Surface charge distribution of the nanostructure at two resonance peaks (590 nm and 760 nm). (B) Schematic of process steps for transfer of the field-enhancing metal-insulator antenna (FEMIA) on a flexible elastomer substrate. (C) SEM cross section view of a 5-layer FEMIA; the inset shows an enlarged view of a single nanostructure. (D) Transferred pattern on the elastomer substrate. The red bounding box marks the region with the pattern. (E) SEM image of the top view of the 5-layer FEMIA on an elastomer.

Based on our previous approach (*26*), we spin coated a layer of NIL resist with a thickness of 350 nm on top of the silicon (Si) wafer, and imprinted the wafer with a pressure of 350 psi at 130 °C. After imprinting, we have removed the residue of the resist using an oxygen (O_2_) plasma at 60 W for two minutes followed by sequential deposition of silver (Ag, 20 nm) and silicon dioxide (SiO_2_, 10 nm) by e-beam evaporation (Figure 2**B(i)**). After deposition, we stripped away the resist in acetone to create a large area (8 mm by 8 mm) of FEMIA nanostructures on Si wafer (Figure 2**B(i)**). The detailed steps are schematically represented in the Supporting Information (Figure **S3**). For transfer of the patterns to flexible substrates, we spin coated polymethylglutarimide (PMGI) on the fabricated nanostructures (Figure 2**B(ii)**) on a germanium coated (Ge) Si wafer, followed by etching of the Ge allowing the resist supported nanostructures to float in a water bath. The floating nanostructures were then scooped from the water bath using a PDMS film (Figure 2**B(iii)**). We used a water-soluble tape to peel off the nanostructures from the PDMS film (Figure 2**B(iv)** & **(v)**). Final transfer to the elastomer was possible by exposing to oxygen plasma and pressing the tape with the nanostructures against the elastomer (Figure **2B(vi)**). The detailed steps for transfer to elastomer is available in the Supporting Information (Figure **S4**). Figure **2C** shows that the patterns are uniform over a large area. Figures **2D** and **2E** show the photograph and SEM image of the pattern on the elastomer, respectively.

Previous studies of transfer printing with irreversible bonding require exposing the nanostructures to oxygen (O_2_) plasma or molecular crosslinking (*27, 28*), hence not suitable for SERS since they tend to oxidize surfaces of Ag reducing the electrical field enhancement (*29*). Here, for the first time, we transfer-printed a large area of Ag based FEMIA nanostructures on an elastomer without compromising e-field enhancement or Raman activity. A key innovation lies in the use of a 100 nm thick germanium (Ge) layer deposited on the silicon wafer, which acts as a sacrificial layer and creates a large area of FEMIA nanostructures on the Ge coated wafer by NIL and multi-layered e-beam evaporation. After lift-off, we have utilized polymethylglutarimide (PMGI) as a carrying film to release the FEMIA nanostructures from the wafer and protect the MIM nanostructures from oxidation during the plasma process. The advantage of PMGI over other polymer films (e.g polymethyl methacrylate) is that we can remove PMGI in an alkaline developer, where organic solvents easily swell any elastomer (*26*).

### Optical Performance of FEMIA

Our normalized experimental reflectance measurement (details in Materials and Methods) closely agrees with the simulation and demonstrates two localized surface plasmon resonance (LSPR) peaks for the 5-layer and 7-layer samples (Figure **3B**). Another important point to note is the array periodicity. When the nanostructures are placed further apart (numerically) the dipolar resonance becomes weaker (Supporting Information figure **S5**). Thus, considering a realistic dimension from the fabrication point of view we have chosen a periodicity of 400 nm in our experiments.

**Figure 3:**
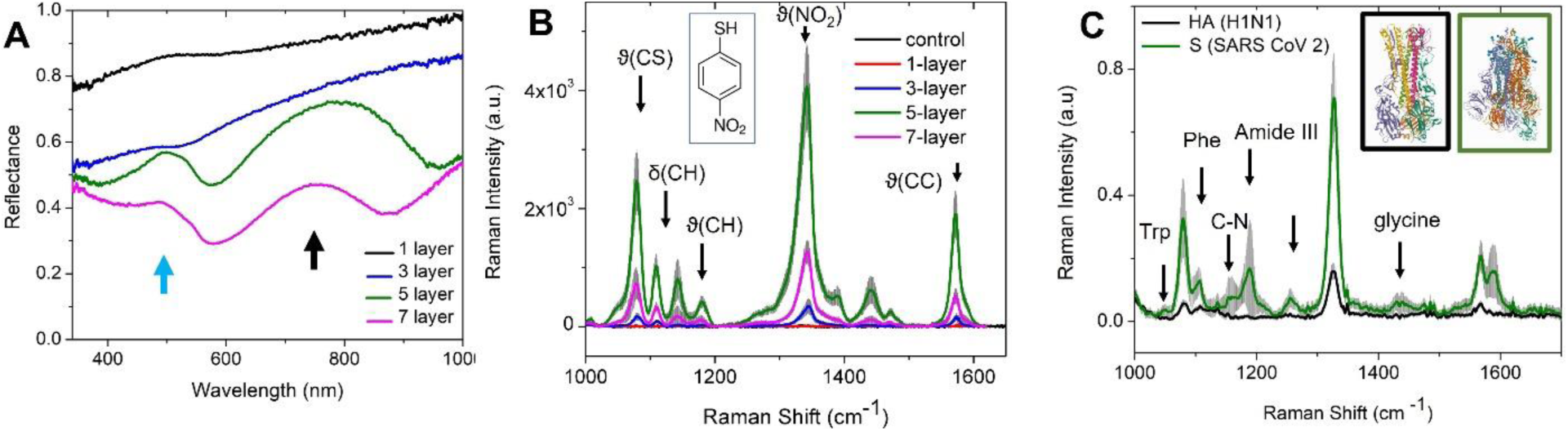
Enhancement characterization of FEIMA. (A) Experimental reflectance spectra of the FEMIA with different layers on silicon. The blue and the black arrow represent the two prominent resonance peaks observed in the 5-layer and 7-layer sample. (B) Raman spectra of 500 µM 4-NTP acquired on different FEMIA substrates and a plain silicon wafer (control). Some of the prominent Raman peaks corresponding to 4-NTP are marked with an arrow. ϑ indicates stretching and δ indicates bending. The inset shows the chemical structure of the molecule. (C) Raman spectra of 0.5 µM spike protein (S) and hemagglutinin protein (HA) found on the surface of SARS-CoV-2 and influenza H1N1 virus, respectively. The prominent peaks are marked in the figure. The inset shows the structures for HA (black box; PDB ID: 1RU7) (*32*) and S protein (green box; PDB ID: 6VSB) (*33*). Trp and Phe refer to tryptophan and phenylalanine. Grey shaded region represents the standard deviation.

The SERS signal enhancing capabilities of the different samples are assessed using 4-nitrothiophenol (4-NTP), a standard Raman reporter. An aqueous solution of 500 µM 4-NTP is dropped on FEMIA substrates with different layers. Raman spectra acquired from the dried drop using a 785 nm laser demonstrate strong signatures of the 4-NTP molecule (*30*). The 5-layer FEMIA, which provides the maximum enhancement (figure **3C)** at 785 nm laser excitation as compared to the other samples, is considered for our study. We have chosen a 785 nm laser for the rest of the study, considering it lies in the tissue transparent region causing minimal damage to biological samples, along with offering a low fluorescence background while maintaining a sufficient signal to noise ratio (*31*). Henceforth unless otherwise stated FEMIA signifies the 5-layer sample.

### Performance of FEMIA for Detection of Antigen

Next, we use the FEMIA substrate to detect purified spike protein (S) and hemagglutinin protein (HA) found on the surface of SARS-CoV-2 and influenza A H1N1 virus, respectively. Vaccines are targeted at the S protein (*34*) and the HA protein (*35*), which are glycoproteins that mediate the attachment of the viruses to the host cell receptors ACE2 and sialic acid, respectively. Thus, identification of the envelope protein is necessary for diagnosis and drug development. Figure **3D** plots the Raman spectra of 0.5 µM of S and HA protein. The S protein sample which is composed of 1288 amino acids as compared to 565 of HA, shows a stronger Raman signature for the same concentration. Various prominent peaks like 1048 cm^-1^, 1138 cm^-1^, 1328 cm^-1^, 1621 cm^-1^(tryptophan) (*36, 37*), 1189 cm^-1^ (amide III) (*38*) 1441 cm^-1^, 1589 cm^-1^(glycine)(*37*) etc. (band assignment in the Supporting Information, table **S1**). Especially prominent in the spectrum of the S protein are the peaks for glycine present in the S2 subunit (*34*) and aromatic amino acid tryptophan at 1048 cm^-1^ (*36*), which has been associated with the coronavirus family.

Although visual inspection provides a moderate level of distinction between the two proteins, to understand the most significant spectral features that enable identification of the purified protein, SERS spectra obtained from the S and HA protein are subjected to principal component analysis (PCA) using Orange data mining software (*39*). Operating without any *a priori* knowledge of the sample composition, PCA projects the spectral data onto a set of linearly uncorrelated (orthogonal) directions (that are called principal components), such that the variance in the original data is captured using only a few PCs (*40*). PC loadings plot (Supporting Information, figure **S6(A)**) of the first 4 PCs shows that the peaks corresponding to amide III, tryptophan and glycine are the important features responsible for the classification of the two proteins, which shows significant clustering in the radial visualization plot (Supporting Information, figure **S6(B)**).

### Classification and Identification of Different RNA Viruses

An enveloped virus is a cocktail of various molecular components like nucleic acid, capsid and proteins. Since the surface protein component could be easily distinguished using SERS signatures on the FEMIA, we next try to see whether viruses with a more complex structure can be distinguished using our platform. In our study, we have considered 4 different viruses - SARS-CoV-2, H1N1 A, which are known to cause respiratory illness and Marburg and Zika virus which cause hemorrhage and fever, respectively. The samples used in our study are viral lysate solutions bought commercially. In order to form the viral lysate, live viruses are allowed to infect cells and are replicated in a culture. The cells are then lysed and the supernatant containing the virus is collected and treated with radiation, heat or chemicals to ensure viral inactivity. In our experiment, drops of lysates with different viruses are allowed to air-dry on the FEMIA substrate before SERS measurement (Figure **4A**). SEM image of a dried drop of the lysate containing SARS-CoV-2 confirms the presence of the virus near the FEMIA nanostructures (Supporting Information Figure **S7**). Zoomed-in image of the region where the analyte is deposited on the substrate shows small particles (either single virus or clusters) in the range of 100 -200 nm consistent with the size range of SARS-CoV-2 (*41*), while they are absent in the region without the analyte (right panel) (Supporting Information Figure **S7**). Figure **4B** plots the SERS spectra collected from the lysates containing the different viruses. The control sample is composed of the cell lysates from the same cell line but without any infection. Although signatures related to protein, nucleic acid and lipid are visible in the spectra, the subtle differences between various samples are indistinguishable by gross visual inspection (band assignment in the Supporting Information Table **S2**). For an unsupervised visual analysis of the entire spectral data set obtained from SERS acquisition on different viral lysates on the FEMIA, the dataset is first subjected to PCA. Figure **4C**, which plots the PC loadings showing various prominent features corresponding to the protein and nucleic acids that differentiate between the various samples enabling clustering of the dataset based on the type of the virus. The PC scores displayed in a multi-dimensional radial visualization plot using Orange data mining software show a clear separation of the spectral data based on the type of virus contained in the lysate (Figure **4D**). It is interesting to note that both SARS-CoV-2 and H1N1 A, which causes similar respiratory illness, occupy nearby clusters. Despite having the base media, the same for all samples, which led to a slight overlap of the cluster corresponding to the control sample with others, a clear separation between various classes is observed. Thus, PCA enables amplifying subtle differences in spectral data.

**Figure 4:**
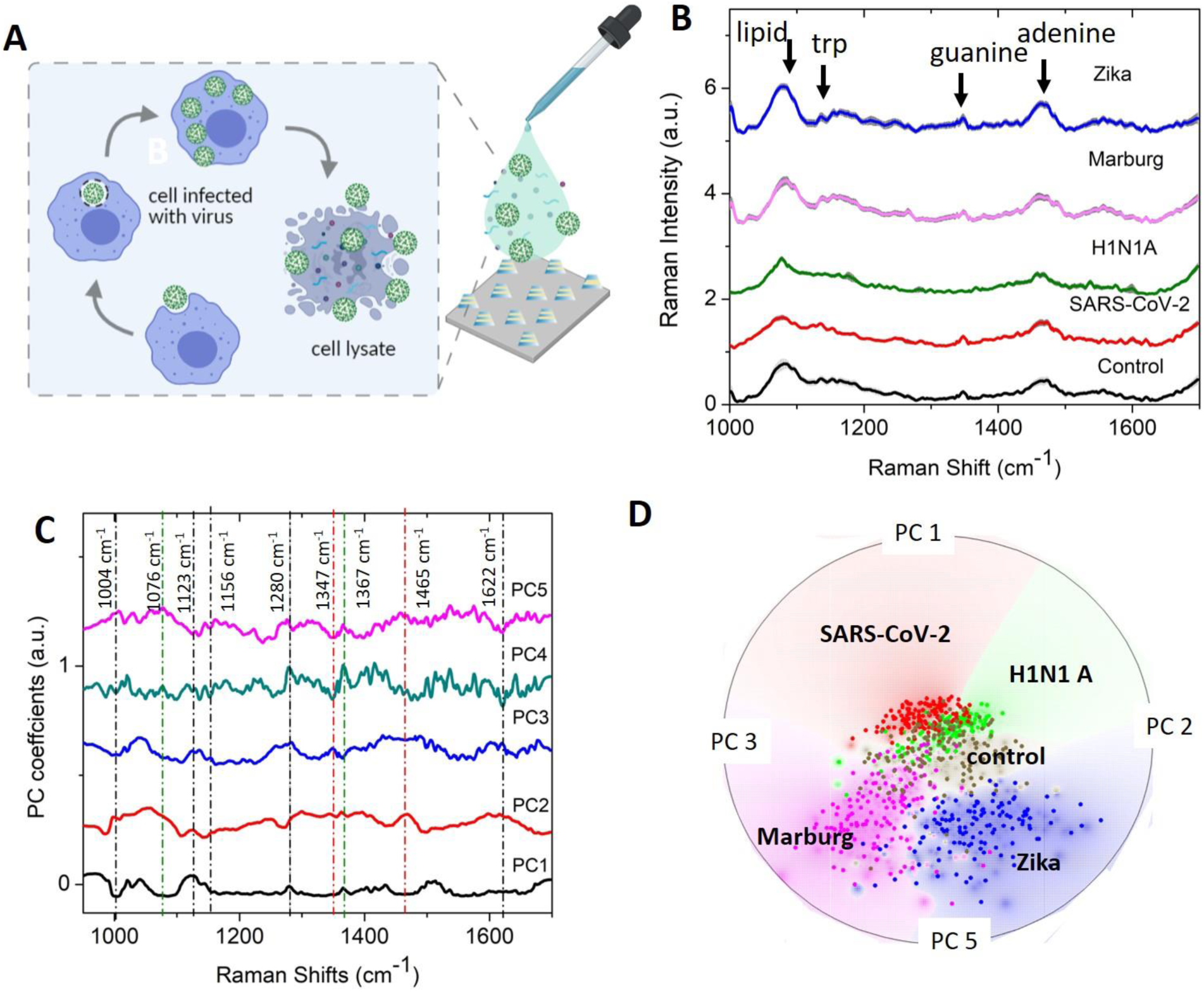
Virus detection on the SERS substrate using unsupervised machine learning. (A) Schematic of the sample details and measurement scheme. Cell lysates infected with the virus are dropped on the FEMIA substrate and allowed to dry. (B) Raman spectra of the cell lysates containing different RNA viruses. The control sample is composed of uninfected cell lysate. Shaded regions represent the standard deviation. (C) PC loadings plot of the entire data set consisting of spectra from the four different viral samples and control. The prominent features are marked with a dotted line-black, green, and red lines indicate the protein, lipid and nucleic acid peaks (E) Radial visualization plot of the PC scores showing data from different samples cluster together. Each dot corresponds to a SERS spectrum.

While principal component-based visualization offers a satisfactory tool for preliminary data exploration, a supervised algorithm is required for the classification and identification of spectral data from an unknown sample. In our study, we employ a random forest algorithm for multiclass classification of the virus spectral dataset. In a random forest classifier, which is based on an ensemble of decision trees, a set of decision trees from a randomly selected subset of the training set is created. The final class of the test object is decided based on votes from the different decision trees (Figure **5A**). For the classification of viral samples, we have trained a multiclass random forest classifier with a training set (randomly chosen two-third of the dataset) composed of 100 decision trees (N). The performance of the model is tested by calculating the out-of-bag classification error on the test dataset (the rest one-third of the dataset) (Supporting Information Figure **S8**). To make the model more robust the random selection of training data and model training and classification are performed 200 times. From the 200 iterations, we observe that the classification accuracy for predicting an unknown viral sample is very high ranging from 83-95% (Figure **5B**). The performance of the classifier is calculated using the confusion matrix (Figure **5C**), which plots the percentage of the test data being assigned to a particular class as compared to its true label. The precision, sensitivity, and specificity for the classification of the SARS-CoV-2 sample are calculated to be 85.4%, 88.77% and 96.1%, respectively. Further, to see whether the accuracy of classification can be improved by grouping the viruses into similar groups, we have performed a binary random forest classification by assigning the dataset related to SARS-CoV-2 and H1N1A to one class (the virus causing respiratory illness) and Marburg and Zika virus to another class (viruses that do not cause respiratory illness). The accuracy for the prediction is 99% (Figure **5D)**.

**Figure 5:**
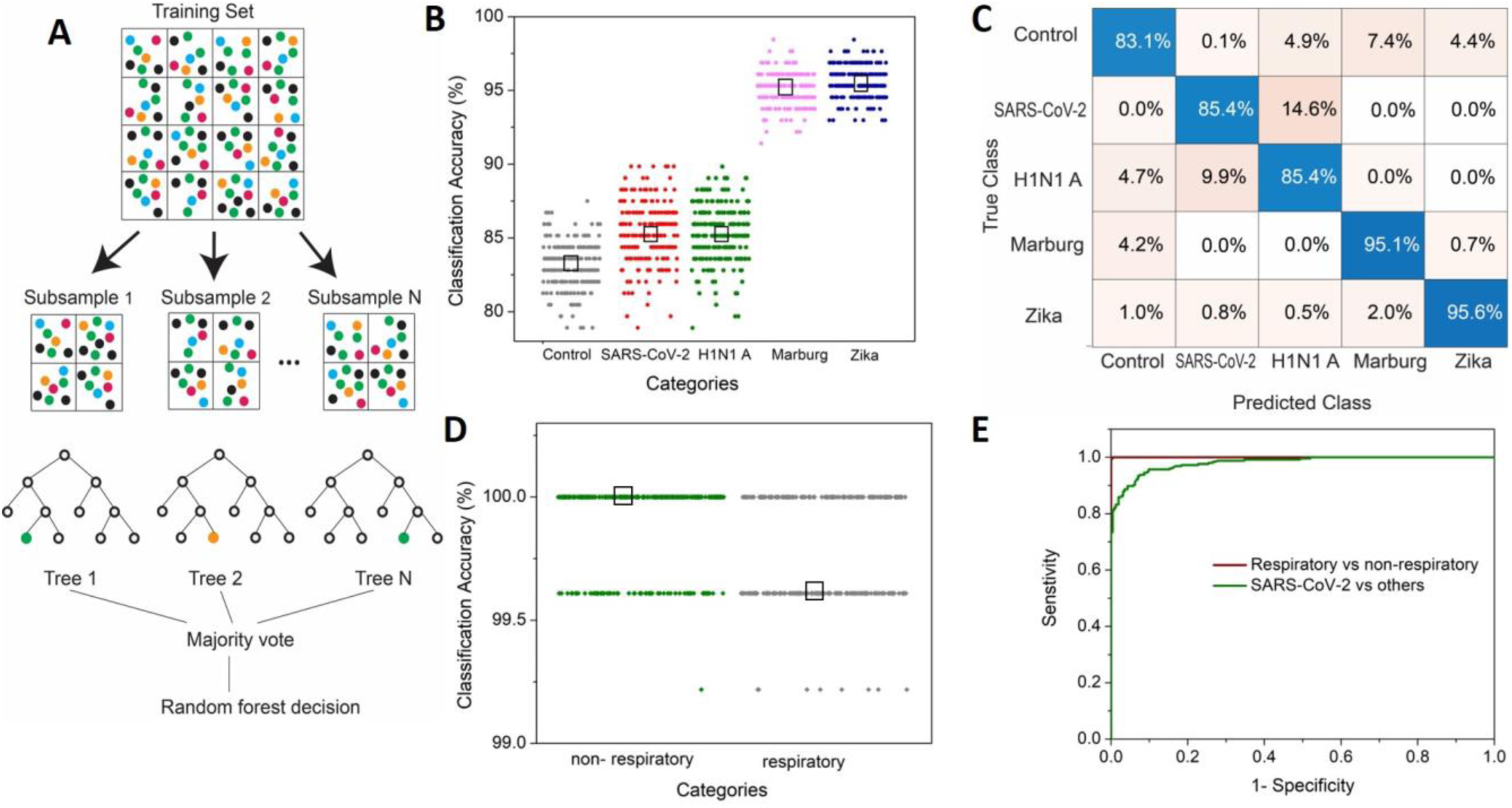
Virus detection on FEIMA using supervised machine learning. (A) Schematic of the random forest algorithm. (B) Classification accuracy for control, SARS-CoV-2, H1N1 A, Marburg and Zika sample using multi-class random forest classifier. The black box represents the median value. (C) Confusion matrix showing the percentage of a sample getting classified into various classes. (D) Classification accuracy using binary random forest classification for SARS-CoV-2 and H1N1 A (respiratory) samples in one class and the other viruses (Zika and Marburg) in the non-respiratory class. (E) ROC curve for the performance of the binary classifications.

To check how well the model performs in predicting datasets related to SARS-CoV-2 in comparison to the other viruses, we employed a binary classification where the dataset related to SARS-CoV-2 is considered in one class as opposed to the rest. We found the classification accuracy to be 92-94% (Supporting Information Figure **S9**). From the receiver operating characteristic (ROC) curve for the two types of binary classification, it is evident that our platform performs very well in distinguishing respiratory vs non-respiratory viruses (area under the curve, AUC -1) as well as SARS-CoV-2 vs other viruses (AUC -0.98) (Figure **5E)**.

### Detection in Pooled Human Saliva on Flexible substrate

Detection of SARS-CoV-2 in the respiratory specimen like nasopharyngeal and oropharyngeal swabs are the current gold standards for diagnostic tests. Other than being relatively invasive this method requires trained operators and close contact with the patient during specimen collection, increasing the risk of transmission. Another method recently being used is detecting the virus in saliva specimens, which can be stored in the ambient till 48 hours and is found to be more suitable for diagnosis of mild and asymptomatic cases (*36, 42, 43*). To check the performance of our platform in detecting SARS-CoV-2 in physiological fluids we spiked pooled human saliva with the cell lysate containing the virus in a 1:1 ratio (Figure **6A**). Our control sample has the saliva spiked with the pristine cell lysate in the same ratio. We dropped the two samples on the FEMIA nanostructures fabricated on an elastomer substrate as discussed earlier. Raman acquisition is performed with lower power and 50x objective to prevent damage to the substrate. Various prominent protein and nucleic acid signatures are visible in the SERS spectra (Figure **6B**) with no clear difference between the two spectra that can be directly derived visually. Saliva is composed of proteins, immunoglobulins, enzymes, mucins, and nitrogenous products making the analyte extremely complex. Thus, we employ random forest analysis to see if the subtle difference in composition due to the presence of the virus can be amplified in such a multi-component matrix. We see that a high classification accuracy of 85-87% is also achieved in this case (Figure **6C**). The ROC for the model is plotted in Figure **6D** with an AUC of 0.94 which implies that the model performs very well in differentiating the virus-containing saliva sample from the control sample. The ability to deploy FEMIA nanostructures and perform COVID detection on flexible substrates points to the possibility of deploying sensors on curved and widely present substrates in the human environment such as fabrics and wearables.

**Figure 6:**
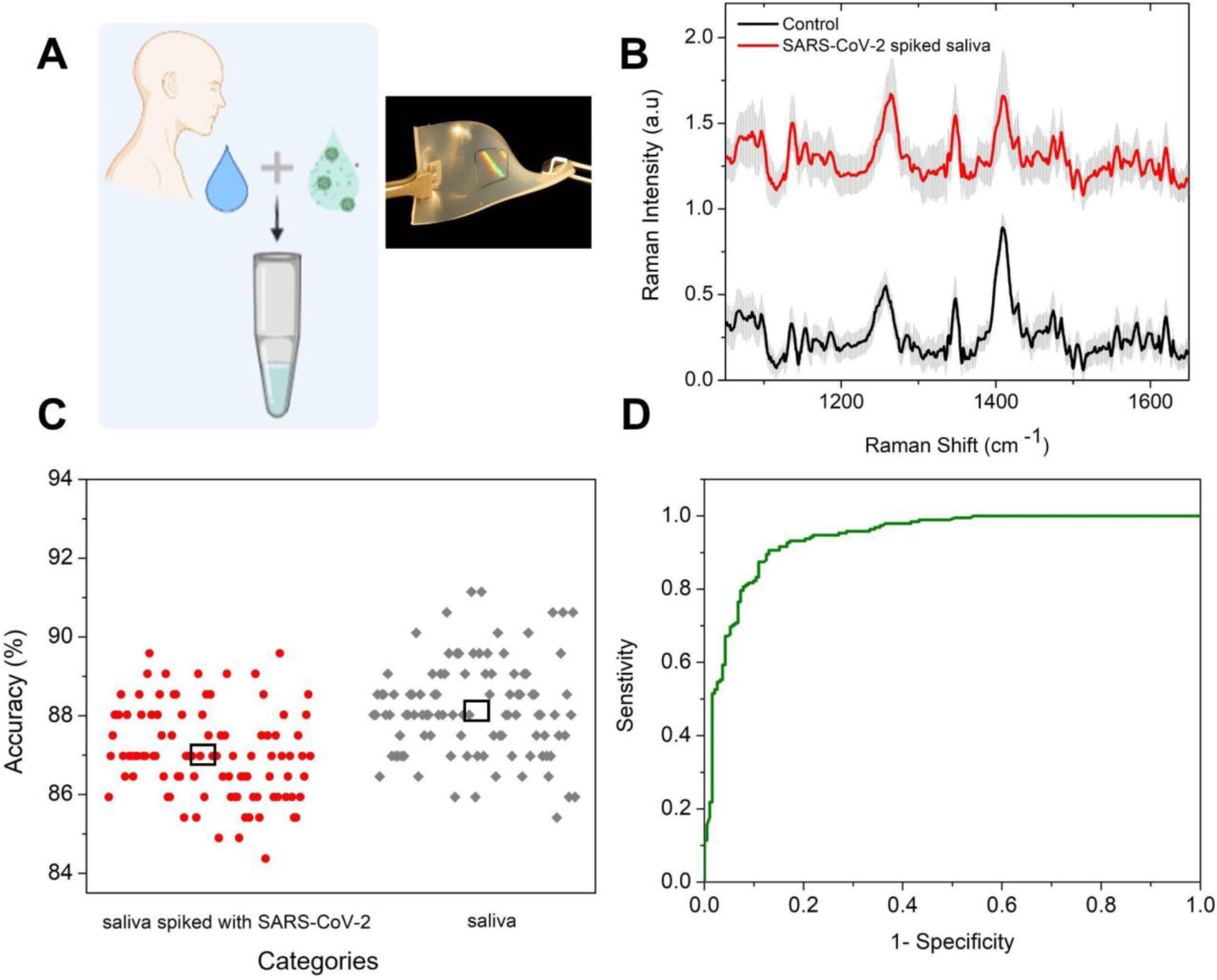
Detection of virus in pooled human saliva on a flexible substrate. (A) Scheme of the experiment. Inset shows a photographic image of the flexible substrate with the nanostructures at the center. (B) Raman spectra of the saliva spiked with SARS-CoV-2 containing cell lysate and the control acquired on FEMIA on a flexible substrate. (C) Classification accuracy using binary random forest classification for the saliva samples. The black box represents the median value. (D) ROC curve for the performance of the model.

## Discussion

One of the most important strategies for predicting and mitigating a pandemic, like the recent COVID-19, is timely and accurate detection and characterization of emerging and reemerging viruses. With the limitations of the current widely used methods, there is a constant need for techniques that are rapid, easy to operate, inexpensive and can be applied to any kind of viruses with no/minimal modifications. The sensing platform we report here has the capability to be built into an inexpensive mass testing technology with a positive result in less than 25 minutes. Although rapid antigen testing can provide marginally faster results (15 mins), the accuracy is lower (*44*).

The large area nano manufactured flexible sensor surface can be mounted on any surface like doorknobs, cylinders, building entrance etc. to enable on-site rapid testing with a handheld Raman setup. The capability of having the entire sensing area on a flexible substrate enables the fabrication of sensor patches that can be placed on face masks or skin for continuous surveillance.

Our choice of nanostructure encompasses an architecture with realistic design parameters to enable one-step lithography-free fabrication while ensuring enough interaction between the nanostructures for increasing the detected signal. It is well known that Raman scattering is a weak phenomenon and signals from biological material are even weaker. Thus, to have a sufficient signal-to-noise ratio, we have used silver nanostructures with resonance matched to the excitation laser frequency for maximizing the Raman scattering signal. A less noisy spectrum enables more accurate use of machine learning algorithms for successfully classifying viruses with similar compositions.

An important thing to note is the requirement of minimal sample preparation in our technique. No chemical or physical pre-processing was required before the measurement. Thus, extensive laboratory training for the operator is not necessary unlike most techniques today. Moreover, a very low sample volume (2.5 µl) with a drying time of fewer than 20 minutes was required. Since no chemical reaction between the analyte and the substrate is involved, the substrate can be suitable for remote collection and storage.

Before using our platform to identify and distinguish different viruses, which are more complex in composition, we checked the performance of our technique in distinguishing purified proteins found on the surface of SARS-CoV-2 and H1N1 A. It was possible to distinguish the signature of the two different proteins directly from the SERS spectra.

For the more complex viral samples, we used supervised algorithms. Random forest classifiers provided a powerful tool to uncover the latent differences in the recorded spectra. The viruses chosen for our study, although being RNA viruses with protein envelope, vary in the composition of the protein and nucleic acid. The subtle differences in the composition, which are reflected as changes in the Raman peak intensities and positions, undetectable visually, can be easily amplified by machine learning. We were able to classify four different viruses with a prediction accuracy of 83% or higher. The viral titer of the SARS-CoV-2 sample (10^9^ copies/ml) used in our experiment is similar to the ones found in patients severely infected with the disease (*45*), indicating that such a technique can be an alternate diagnostic scheme.

Our sensing platform also performed well in multicomponent media like the pooled saliva. We spike pooled human saliva with SARS-CoV-2 viral samples and show that detection is also possible using our method in physiological fluids in a non-invasive manner. Although the acquired spectra are relatively noisier due to the interference from various components of the complex matrix, the random forest classifier was able to differentiate based on the subtle difference due to the presence of the virus.

A drawback of our sensing platform is the requirement of known samples for training the algorithm. Also, a better prediction accuracy to minimize false negatives and positives requires a larger dataset, which will increase the requirement for computational resources.

Finally, for in-field applications like mass testing in airports, shopping malls, etc., our platform can be paired with a hand-held Raman spectrometer. The manufacturing cost of the sensor can be estimated to be tens of dollars per chip, which can be reduced by scaling up.

In summary, we have developed a scalable, flexible, plasmonically active SERS substrate for fast identification of viruses like SARS-CoV-2, H1N1 A among others. Raman spectroscopic sensors combined with machine learning offer the possibility for rapid label-free testing without labeling of biomolecules and with a high degree of accuracy. We envision that this platform when combined with a portable Raman spectrometer could be utilized to create a sensing device for rapid and on-site mass testing of viruses and pathogens.

## Materials and Methods

### Fabrication of FEMIA arrays on a rigid (silicon wafer) substrate

We fabricated the array of metal insulator metal (MIM) nanostructures by nanoimprint lithography (NIL). Briefly, we spin-coated a layer of the NIL resist (mr-I 7030, micro resist technology) with a thickness of 350 nm on top of a silicon wafer. We imprinted the nanopatterns using a commercial low-cost Si master stamp (LightSmyth grating) and a Nanonex Advanced Nanoimprint Tool NX-B200. After imprinting, we stripped away the residue of the resist using an oxygen (O_2_) plasma at 60 W for 2 minutes. Our MIM nanostructures have multiple i.e 1, 3, 5 or 7 MIM layers. Each layer has silver (Ag, 20 nm), silicon dioxide (SiO_2_, 10 nm) and silver (Ag, 20nm) which were deposited by e-beam evaporation. We also deposited 2 nm of chromium (Cr) between the silicon wafer and Ag and in between each layer of Ag and SiO_2_ and SiO_2_ and Ag as well as to improve adhesion. After deposition, we sonicated the sample in acetone to dissolve the mr-I 7030 and generate arrays of MIM nanostructures on top of the Si wafer.

### Fabrication of FEMIA arrays on a flexible (elastomer) substrate

We thermally evaporated a thin film of germanium (Ge) with a thickness of 100 nm on a Si wafer as the sacrificial layer. We created arrays of multiple MIM nanostructures on top of the Ge coated wafer using the same approach described in the previous section. We spin coated a layer of polymethylglutarimide (PMGI SF6, Kayaku Advanced Materials) as a carrying film to transfer the arrays of MIM nanostructures. We etched away the Ge sacrificial layer in water and picked up the film using a mm-scale thick piece of PDMS. After the film is completely dried, we peeled off the film from the PDMS using water-soluble tape (3M 5414). We activated the surfaces of the bottom of the MIM nanostructures (SiO_2_) and elastomer (Dragon Skin™) by O_2_ plasma. We bonded the MIM and elastomer by pressing both surfaces together and removing the tape and PMGI in water and alkaline developer (MF 26A, Kayaku Advanced Materials).

### Simulation details

For the numerical simulation, we have used finite element analysis (COMSOL Inc., Burlington, MA, USA). We have used a periodic boundary condition to simulate infinitely repeating FEMIA nanostructures in 2 dimensions. We have considered a constant dielectric constant value of 2.09 for the silica layer and a frequency-dependent dielectric constant for the silver layer (*46*). We have considered dynamic meshing where the maximum and minimum size for the FEMIA nanostructure is 50 nm and 0.5 nm respectively and λ/6 (λ - wavelength of incident light) for the rest. The boundary conditions at the input and output ports were assigned as a perfectly matched layer. The dimensions for the FEMIA were considered as measured in SEM, i.e, the thickness of silver and silica were 20 nm and 10 nm respectively. We considered the shape of the nanostructure as conical (with rounded edges) having a bottom radius of 130 nm and center to center distance between adjacent nanostructures was 660 nm.

### Reflectance measurement

Reflection spectra were acquired by a USB 4000 spectrometer (Ocean Optics) and a tungsten halogen lamp (LS-1) (Ocean Optics). The spectra were averaged over 3 measurements and normalized.

### Raman measurement

We used an XploRA PLUS Raman microscope (HORIBA Instruments Inc., Edison, NJ, USA) for the Raman measurements. For the 4-NTP study, the Raman scattered signal was acquired through a 100x Olympus objective for 1 s and averaged 3 times, 10% power. For the purified protein study, a 100x Olympus objective was used for 0.5 s and averaged 20 times, 25% power. For each analyte, signals were collected from 64 different spots on two different substrates and averaged.

For the viral samples, the acquisition parameters were: 100x Olympus objective, 0.5 s acquisition time, 30 accumulations, 25% power, 128 spectra.

For the saliva samples: 50x Olympus objective, 0.5 s acquisition time, 30 accumulation, 1 µW 25% power, 128 spectra.

For all the samples 2.5 µl of the solution was used. A thermoelectrically cooled charge-coupled device camera (1024 × 256OE Syncerity, HORIBA Instruments Inc., Edison, NJ, USA) collects the scattered spectra. We have calibrated the Raman shift with silicon.

### Samples details

4-NTP was bought from Sigma-Aldrich, St. Louis, MO.

HA protein was bought from HA protein from Sino Biological A/WSN/1933 (Cat# 11692-V08H), S protein BEI Resources

Following virus samples and control cell lysates were bought from BEI Resources

Influenza A Virus, A/New York/18/2009 (H1N1) (10^7^ copies/ml)

SARS-Related Coronavirus 2 (*47*), Isolate USA-WA1/2020 (10^9^ copies/ml)

Zika virus, PRVABC59 (10^10^ copies/ml)

Marburg Marburgvirus, H. sapiens-tc/AGO/2005/Angola-200501379 (10^11^ copies/ml)

All samples have cell lysate from *Cercopithecus aethiops* kidney epithelial cells (Vero E6) as the base media.

Control - Vero 6 cell lysate from BEI Resources

Human pooled saliva - Innovative Research

The virus samples were handled in BSL-2 laboratory conditions. After the measurement the samples were discarded in a 10% bleach solution.

### Data Analysis

The Raman spectra collected were subjected to a background subtraction via a polynomial fitting algorithm (*48*). We have applied a 5^th^ Savitzky Golay filter with frame length 21 on the resultant spectra. This was followed by peak normalization with the maximum intensity value. We used Matlab 2019b for the spectral pre-processing.

TP - true positive, TN - true negative, FP - false positive, FN - false negative.

Precision: TP/(TP+FP); Recall: TP/(TP+FN); Specificity: TN/(TN+FP).

### PCA

The pre-processed data were subjected to PCA using Orange data mining software (*39*). Operating without any a priori knowledge of the sample composition, PCA projects the spectral data onto a set of linearly uncorrelated (orthogonal) directions (that are called principal components), such that the variance in the original data is captured using only a few PCs. The radial visualization plot maps the PC scores onto a two-dimensional space to permit analysis of inter-vs intra-class variance of the different analytes. We employed the VizRank algorithm in the Orange data mining software to select the PCs based on their ability to obtain informative projections of the class-labeled data.

### Random Forest Analysis

For random forest analysis, the number of spectra was made equal for all classes. This equalization was done by first getting a minimum number of spectra from each class and then by randomly picking a minimum number of spectra from other classes. This method ensured that the random forest results are not biased to any class. Thereafter, the TreeBagger built-in Matlab function was used to perform classification-based random forest with sample replacement and curvature-based predictor selection. After that out-of-bag prediction was used to predict classes. Since the initial spectra selection from different classes were at random, we repeated the analysis 200 times and the final classification results included all the iterations.

## Data Availability

All data produced in the present study are available upon reasonable request to the authors

## Acknowledgement

Spike protein was produced under HHSN272201400008C and obtained through BEI Resources, NIAID, NIH: Spike Glycoprotein (Stabilized) from SARS-Related Coronavirus 2, Wuhan-Hu-1 with C-Terminal Histidine Tag, Recombinant from HEK293F Cells, NR-52397. The H1N1 A was obtained through BEI Resources, NIAID, NIH: Influenza A Virus, A/New York/18/2009 (H1N1)pdm09, BPL-Inactivated, NR-49451. The Marburg virus was obtained from BEI Resources, NIAID, NIH: Marburg Marburgvirus Prototype Isolate Marburg Virus/H. sapiens-tc/AGO/2005/Angola-200501379, Infected Cell Lysate, Gamma-Irradiated, NR-50544. The Zika virus sample was obtained through BEI Resources, NIAID, NIH: Zika Virus, PRVABC59, Heat-Inactivated (Lyophilized), NR-50432. Vero E6 Cell Lysate Control was obtained through BEI Resources, NIAID, NIH: Vero E6 Cell Lysate Control, Gamma-Irradiated, NR-53258. We acknowledge funding from the National Science Foundation CMMI-2033349. D.H.G. would like to dedicate this paper to his father Abel Gracias who passed away from complications due to COVID in Jan 2021.

