## supporting information for "Label-Free SARS-CoV-2 Detection on Flexible Substrates"

Ishan Barman

Johns Hopkins University

Whiting School of Engineering

Department of Mechanical Engineering

Latrobe Hall 103

Baltimore, MD 21218, USA

### Details of the Finite Element Electromagnetic Simulation Model

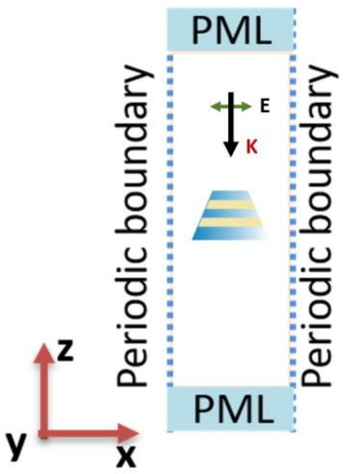


**Figure S1: Scheme of simulation.** The direction of polarization and the propagation of the incident field is shown by the green and black arrow, respectively.

### Absorption characteristics of the FEMIA

From the absorption coefficient an approximate location of the plasmon resonance for the FEMIA with different number of layers can be identified.

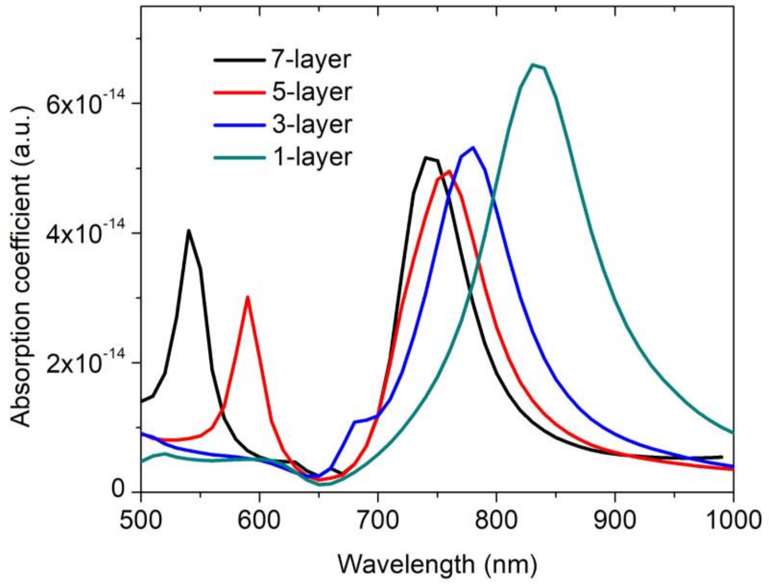


**Figure S2:** Absorption of a 2-dimensional array of FEMIA nanostructures with a single layer of silver (1 layer), 1 layer of silica sandwiched between 2 layers of silver (3 layer), 2 layer of silica sandwiched between 3 layers of silver with layers alternating (5 layer), 3 layer of silica sandwiched between 4 layers of silver with layers alternating (7 layer).

### FEMIA fabrication on rigid (Si) substrate

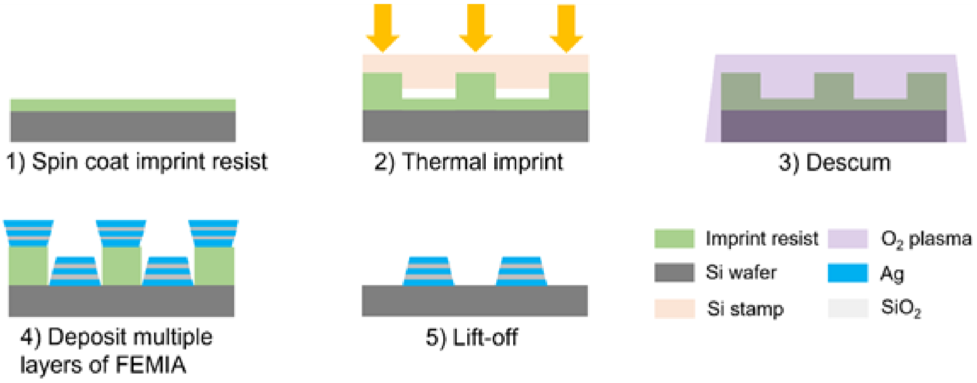


**Figure S3:** Schematic of the detailed steps for fabrication of FEMIA nanostructures on Si wafer.

### FEMIA transfer on flexible substrate

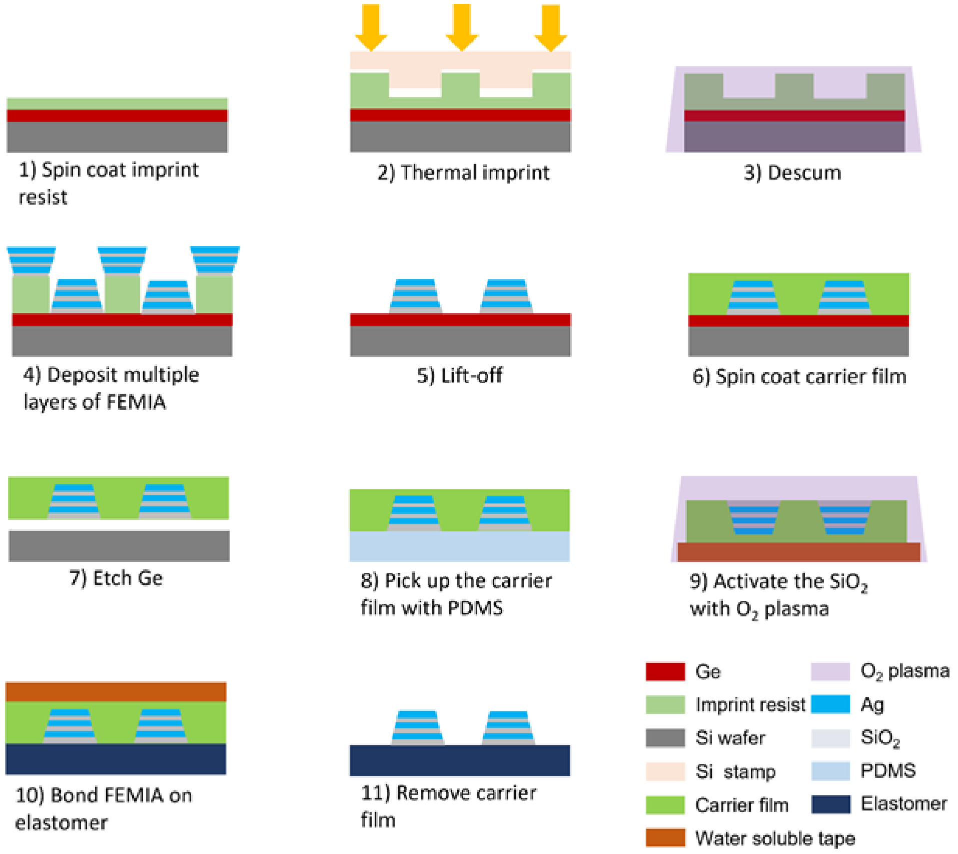


**Figure S4:** Schematic of the detailed steps for transfer of FEMIA nanostructures on flexible substrates

### Dependence of plasmon resonance on array periodicity

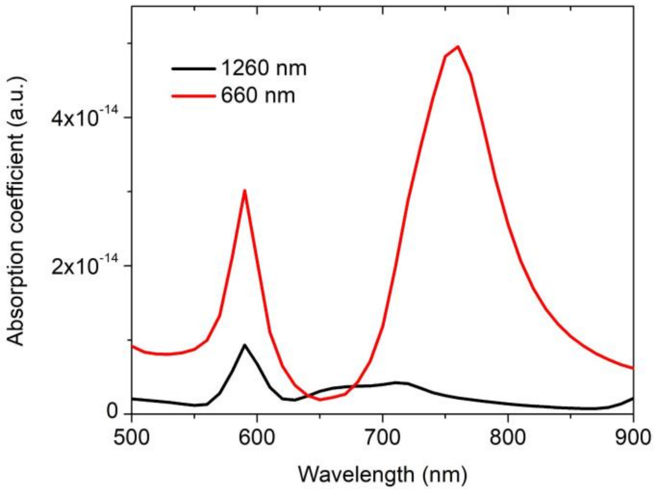


**Figure S5:** Absorption of an infinite 2-dimensional array of 5-layer FEMIA for different array periodicities, i.e. centre to centre distance of 1260 nm and 660 nm.

**Table S1:** Peak assignment of the prominent peaks in the Raman spectra of HA and S protein

| **Raman shift ($cm^{-1}$)** | **Assignment** |
|---|---|
| 1048, 1138, 1328, 1621 | Tryptophan |
| 1104 | Phenylalanine |
| 1155 | C-N stretching |
| 1189, 1258 | Amide III |
| 1441, 1589 | Glycine |
| 1453 | C-H stretching of glycoprotein |
| 1567 | valine |

### PCA on SERS spectra of HA and S protein

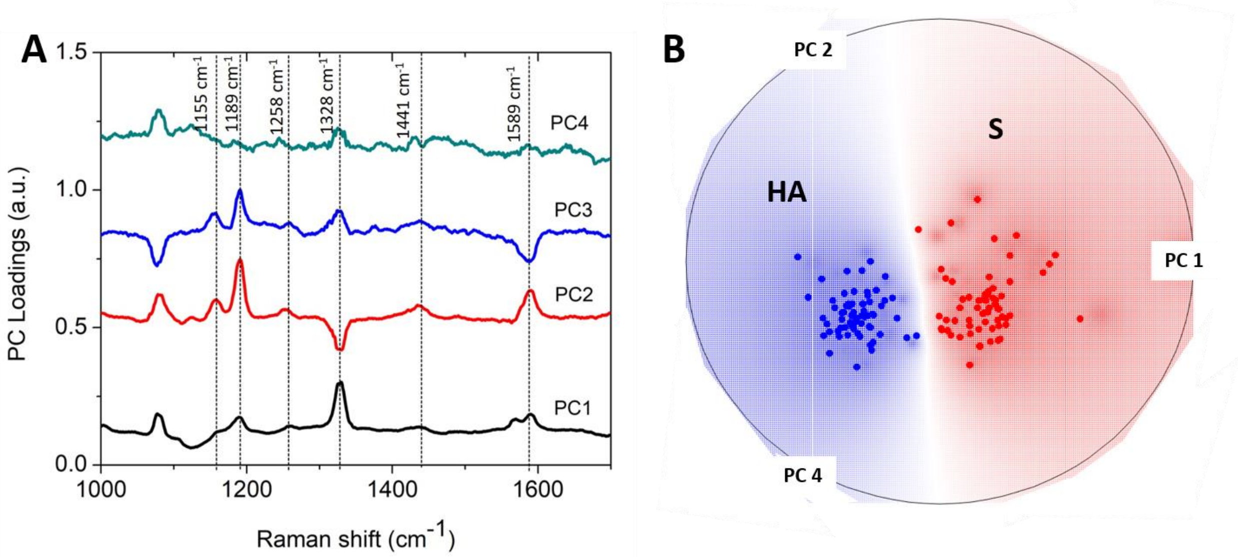


**Figure S6:** (A) The first four principal components obtained from the entire date set of HA and S protein. The prominent features are represented by the dotted lines. (B) Radial visualization plot of the most relevant PC scores demonstrating the clustering of the two types of proteins.

### Dried drop of viral lysate on FEMIA

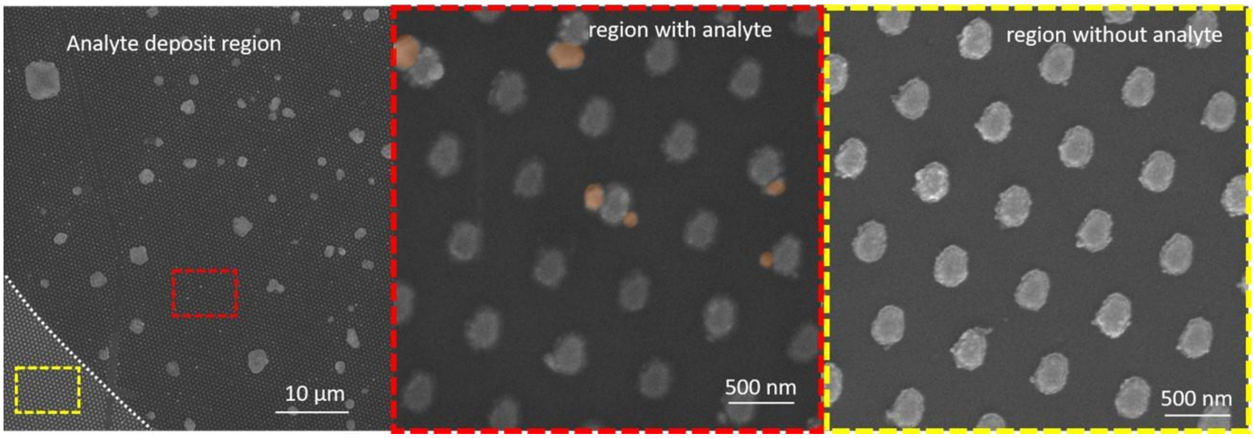


**Figure S7:** SEM image showing a dried drop of cell lysate infected with SARS-CoV-2 (left panel) on the FEMIA. Zoomed in image of the region with the analyte (middle panel) and without the analyte (right panel). The virus particles are false colored orange for better visualization.

**Table S2:** Peak assignment of the prominent peaks in the Raman spectra of the cell lysates containing the virus

| **Raman shift ($cm^{-1}$)** | **Assignment** |
|---|---|
| 1004 | Phenylalanine |
| 1076 | lipids |
| 1123 | C-N (protein) |
| 1138, 1622 | tryptophan |
| 1156 | C-N (protein) |
| 1246, 1280 | Amide III |
| 1347 | guanine |
| 1367 | lipid |
| 1465 | adenine |

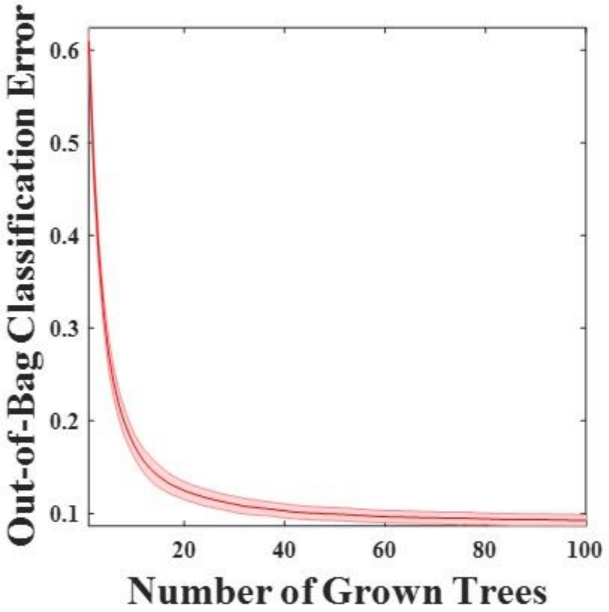


**Figure S8:** Evaluation of classification error vs number of decision trees, shows that the error falls below 10 % with the inclusion of 100 trees.

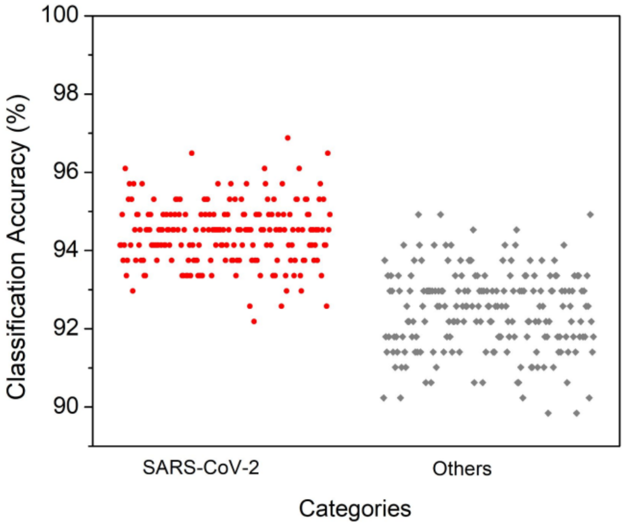


**Figure S9:** Classification accuracy for binary random forest classification with SARS-CoV-2 sample in one class and the other viruses (H1N1 A, Zika and Marburg) in the other class.
